# A survey assessing antimicrobial prescribing at UNRWA primary health care centers in Jordan

**DOI:** 10.1101/2022.01.14.22269315

**Authors:** ESF Orubu, S Albeik, C Ching, R Hussein, A Mousa, M Horino, R Naqa, M Elayyan, R Saadeh, MH Zaman

**Affiliations:** Social Innovation on Drug Resistance, Boston University, Boston, United States of America; Department of Biomedical Engineering, College of Engineering, Boston University, Boston, United States of America; United Nations Relief and Works Agency for Palestinian Refugees in the Near East, UNRWA, Jordan

**Keywords:** antimicrobial resistance, stewardship, humanitarian, primary health care, UNRWA

## Abstract

Antimicrobial resistance (AMR) is a public health emergency. There is insufficient information on AMR in the context of humanitarian settings. An understanding of behavioural and institutional level factors can strengthen antimicrobial stewardship. This study used a semi-structured questionnaire to assess both knowledge, attitudes and practices (KAP) on antimicrobial use, resistance and stewardship, and options to improving prescribing, among prescribers at the Primary Healthcare facilities of the UNRWA Jordan field office. Responses to the KAP questions were evaluated using the Capability, Opportunity, Motivation, Behaviour (COM-B) framework and Bloom’s cutoffs. For each framework component, Blooms cutoffs and interpretations were: >80%, “good”; 60-79%, “moderate”; and <60%’ “poor”. Fourteen options to improve prescribing were each assessed using 5-point Likert scales from very unhelpful to very helpful, aggregated by helpful and very helpful and ranked as: >90%, best/most acceptable; >80-90%, as acceptable; and, 70-80% as maybe acceptable/good. The questionnaire response rate was 59% (37/63) with a completion rate of 92% (34/37). Aggregate scores for real knowledge on AMR was 97%; opportunity to improve prescribing 88%; and motivation 16% - participants did not believe that there was a connection between their prescribing and AMR or that they had a key role in helping control AMR. Good options (74% aggregate score) to improving prescribing were the availability of guidelines and resistance data. There was good knowledge of AMR and good opportunities, but poor motivation for rational prescribing or behavioral change. There is a clinical need for antimicrobial resistance data to promote rational antibiotic prescribing.

## Introduction

Antimicrobial resistance (AMR), an inherent or acquired ability of disease-causing microorganisms to survive therapeutic doses of antimicrobials is a pressing global health challenge ^1,2^. Anthropogenic factors including the non-evidence-based prescribing, dispensing and use of antimicrobials including for non-therapeutic indications in livestock, the use of substandard or falsified antimicrobials, poor adherence in patients and environmental (metal and) antibiotic run-offs accelerate AMR ^3^. Low- and Middle-Income Countries (LMICs) are predicted to bear the greater burden of AMR ^4^. Generally, communities living in cramped environments with limited resources are at a high risk of developing drug resistant infections ^3^. This may be particularly true for refugees and the forcibly displaced, where geo-socio-economic determinants predisposing to disease combine with sub-optimal access and the excessive use of antibiotics ^5–7^.

Jordan is one of the world’s largest hosts for displaced populations. In 2020, it was the ranked the 10^th^ largest host country for refugees and the internally displaced ^8^. Jordan, historically, also has the highest number – over 2.3 million – of registered Palestine refugees, (PRs) under the United Nations Relief and Works Agency for Palestine Refugees in the Near East (UNRWA) program among the five fields of its operations (Gaza, the West bank, Syria, Lebanon, and Jordan). In Jordan, UNRWA provides free health care services (including outpatient services) to Palestine refugees through a network of 25 primary health care centers. There are scant studies on antibiotic utilization among PRs settlements in Jordan. One previous study from 2018 on the knowledge, attitude and behavior of PRs regarding antibiotics surveyed 250 adults at 4 different UNRWA health centers (HCs) in Jordan (Irbid, South Baqaa, Taybeh and Marka) ^9^. The study revealed a poor understanding of antibiotics. Self-medication, over-the-counter (OTC) purchase as well as using left-over antibiotics were commons form of inappropriate antibiotics use. Further, there was a knowledge gap in the correct use, and of side effects from irrational use, of antibiotics among participants. Long waiting times drive the OTC purchase of antibiotics, even though there is high trust in doctors among patients in refugee camps ^9^. This data suggests that the risk for AMR among refugees, arising from misuse of antibiotics, is high. The study, however, did not identify which specific antibiotics were most commonly used and did not consider the prescribing and dispensing practices of healthcare workers and pharmacists. Thus, there are gaps in our understanding.

This study, as part of a series assessing refugees’ access to quality healthcare services and AMR, performs an evaluation within the context of a primary health care system. The aim was to assess and analyze antimicrobial prescribing in the Jordanian UNRWA sites. The objectives of the study were two-fold: (i) describe antibiotic prescribing patterns, as proxies of use (ii) assess knowledge, attitude, and practices on antimicrobial resistance and use, with a view to understanding barriers to antimicrobial stewardship.

## Methods

### Ethics

Ethics application was reviewed by the Institutional Review Board of Boston University and granted exemption (5681X). Ethical approval was also obtained from UNRWA’s research review board before questionnaire administration.

### Study design

Cross-sectional electronic survey design. This study adopted the Checklist for Reporting Results of Internet E-Surveys (CHEERIES) protocol for questionnaire development and reporting ^10^.

### Study setting

Primary Health Care (PHC) facilities run by the UNRWA in Jordan. A convenience sample of 16 PHCs were selected based on size and accessibility ^11^.

### Participants’ inclusion and exclusion criteria

Only medical officers, gynecologists, and dental surgeons working at the selected UNRWA PHCs who prescribe antimicrobials were included in the survey. All other healthcare workers at the selected PHCs and all healthcare workers outside these facilities were excluded.

### Study instrument

The study instrument was a semi-structured questionnaire based primarily on a previously validated knowledge, attitude, and practice survey used in Europe ^12^. This was complemented by other previously used survey instruments ^13,14^ further adapted to a humanitarian setting by including questions assessing prescribers’ perceptions of AMR in the forcibly displaced ^15^.

The questionnaire comprised 4 sections containing 43 questions assessing demographics, current prescribing behavior, and potential for behavioral change (Table 1 and Supplement 1). Section I had 8 questions on demographics. Section II included 16 questions to describe current prescribing practices and antibiotic prescribing patterns. Section III contained 16 questions assessing knowledge and attitudes. These questions were structured using the Capability, Opportunity and Motivation for behavioral change (COM-B) framework. Capability assessed prescribers’ knowledge, awareness and perceptions or attitudes. In this study, we described capacity as consisting of decision making, knowledge, awareness/perceptions, attitudes, and infection prevention and control measures (hand hygiene). Knowledge was both real and assumed. Opportunity included perceptions about both individual- and facility-level factors that make for rational prescribing. Motivation was self-held beliefs about influence on rational prescribing and association between prescribing behavior and AMR. Section IV contained 1 question with 14 options to improve antimicrobial prescribing. There were also 1 introductory question on consent and 1 final question on additional comments.

**Table 1.**
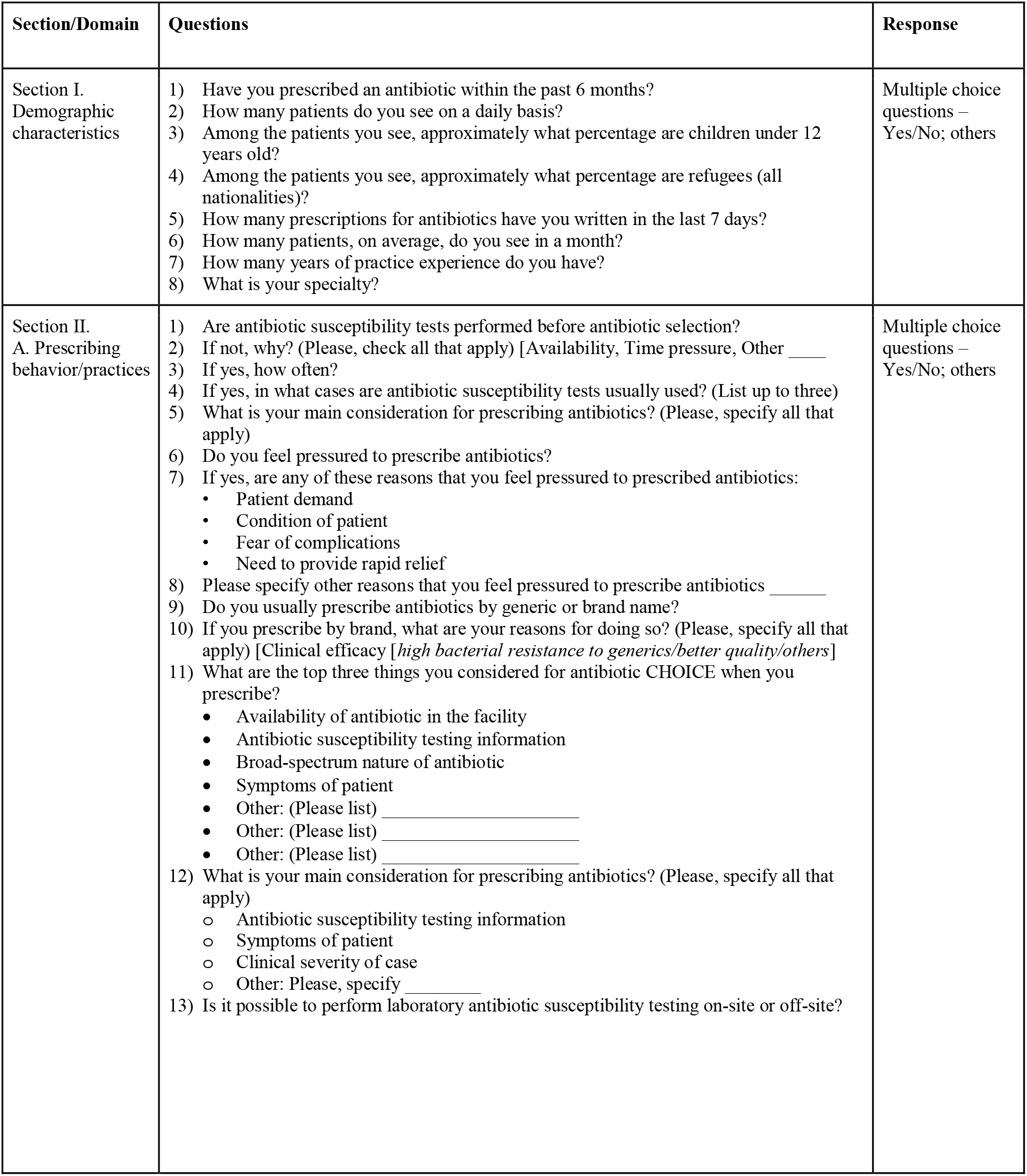
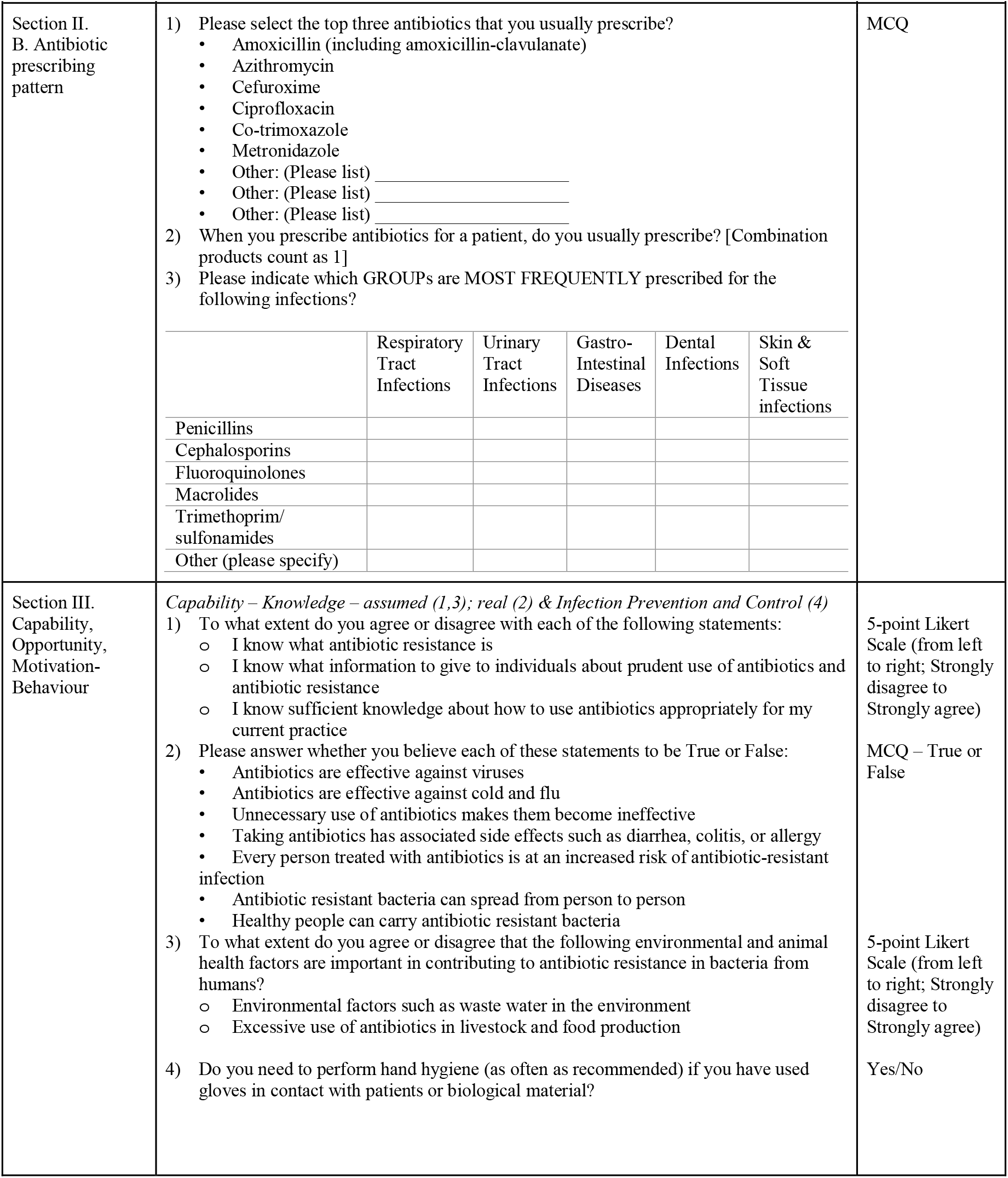
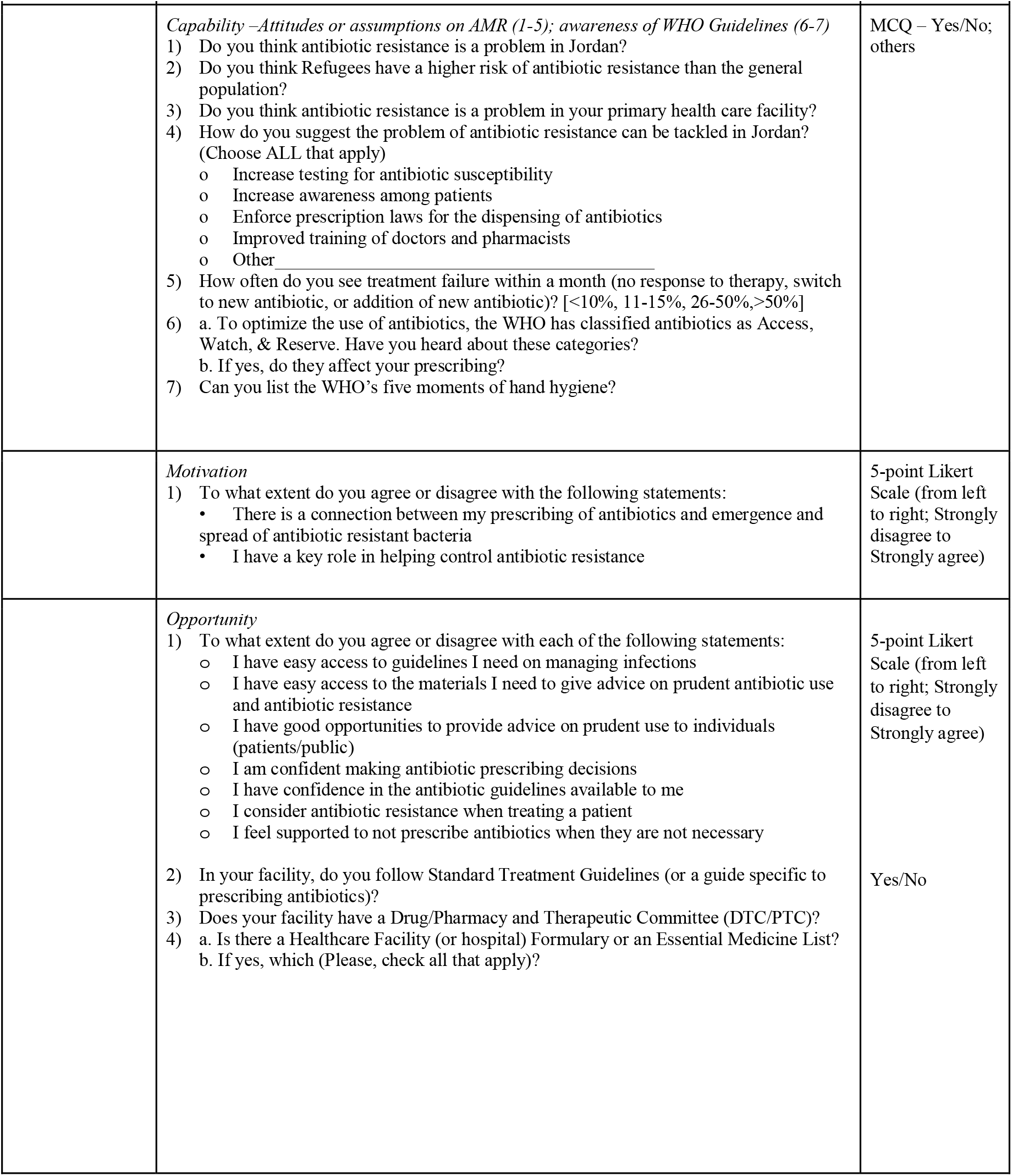
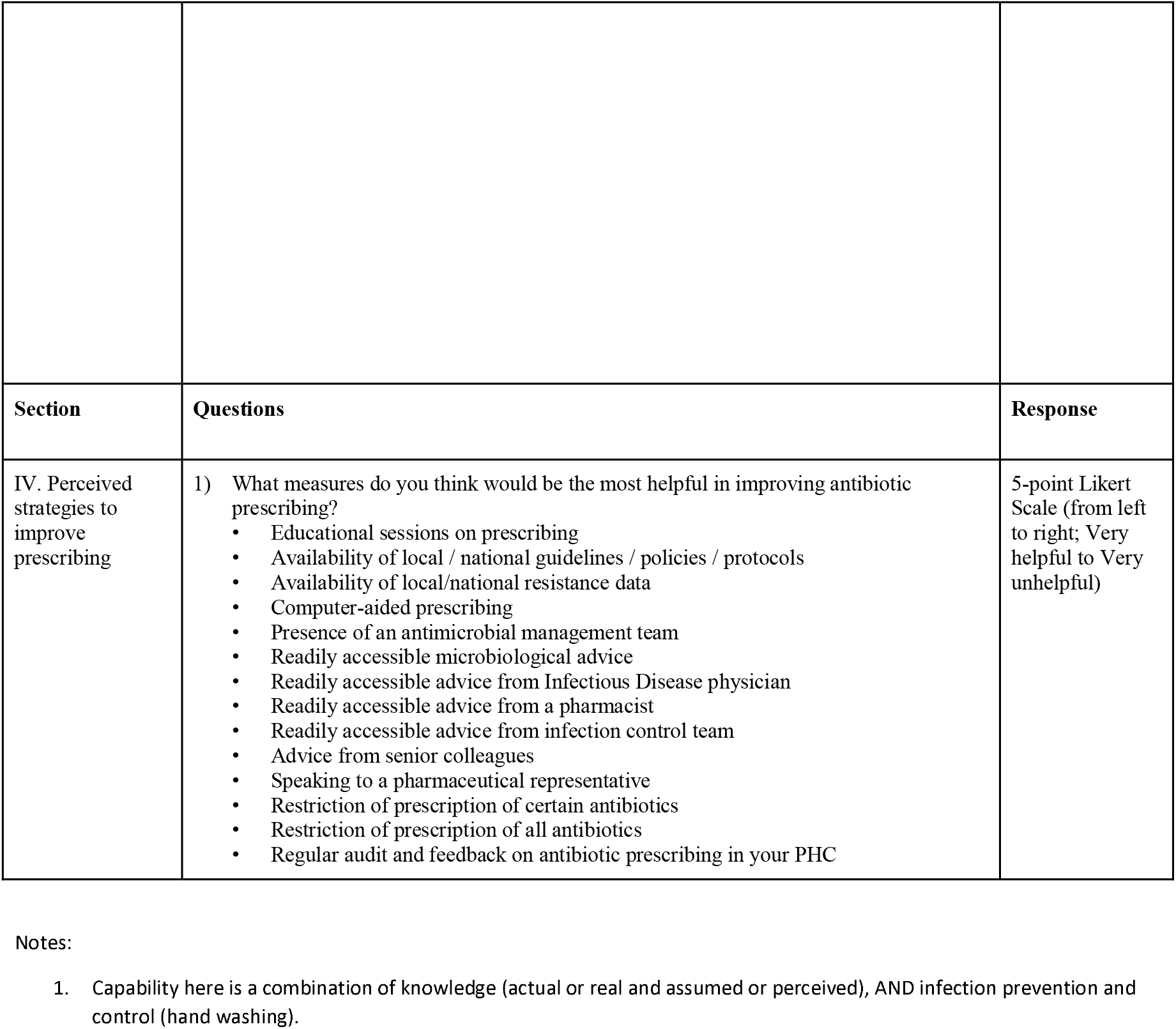
Interview guide for the knowledge, attitudes, and practices survey of prescribers at UNRWA PHC, Jordan showing sections, questions, and response types. Section I collected the characteristics of clientele and prescribers. Section II describes current prescribing behavior and practices. Section III assessed knowledge and attitudes on rational prescribing mapped onto the COM-B framework.

Items in the questionnaires were randomized. Using adaptive questioning, certain questions were only required to be answered based on whether a preceding question was answered. To ensure completeness when filling the questionnaires, some questions were pre-specified and made mandatory, where these questions had to be filled before the questionnaire could be submitted. The questionnaires were made available in English and Arabic. The Arabic version was provided by SH and verified by SA. Both versions incorporated an introductory information and consent section.

### Piloting

The questionnaire was piloted in a PHC not among those selected to be sampled. Based on the results from this pilot, the instrument was modified to ensure validity in the study locations. Modifications included changes to the Arabic translation, a consent section with termination where consent was not granted, and the review step to ensure completeness.

### Sample size

The sample size was all consenting physicians out of a list of 63 prescribers provided by Jordan field office at the 16 selected PHCs.

### Questionnaire administration

Questionnaires were self-administered. Participants were contacted through a list of emails and/or WhatsApp numbers for all medical officers currently working in the selected 16 PHCs in Jordan. The questionnaires were shared (as links) with the participants. Consent was obtained from all participants by ensuring that only those who consented continued to fill the questionnaire. Questionnaires were administered from August 1 to September 30, 2021.

### Data analysis

Data analysis was descriptive with responses expressed as proportions (%). Current prescribing practices were described. Aggregate scores across participants in the capability (real knowledge), opportunity and motivation components of behavior were computed and evaluated against Bloom’s cutoffs. For real knowledge, opportunity and motivation a mean aggregate score >80% was assessed as “good”, 60-79% as “moderate”, and <60% as “poor” ^16^. The perceived options to improve antibiotic prescribing were aggregated and ranked. On the 5-point Likert Scale, “very helpful” and “helpful” were aggregated into the new dummy variable “helpful” and neutral, unhelpful and very unhelpful aggregated into a new “unhelpful” dummy variable. The aggregated scores under the new “helpful” category for each of the 14 options to improve prescribing were then ranked with aggregate scores >90% assessed as the best, or most acceptable, option(s); >80-90%, as acceptable; and, 70-80% as maybe acceptable, or good.

## Results

### Characteristics: response rate and participants demographics

The questionnaire response, or participation, rate was 59% (37/63). However, the completion rate, or participants consenting to, and completing, the survey was 92% (34/37).

Participants were mostly general medicine specialists (75%, 25/34). The majority had more than 6 years’ experience (35% with 6-10 years’ experience; 38% with 11-15; and 9% with >15 years).

The complete demographic characteristics and detailed results are included in the supplement.

### Antibiotic prescribing practices

All respondents had prescribed antibiotics in the 6 months preceding the study. All prescriptions were by generic names (100%, 34/34). Prescribers (91%, 31/34) reported the most frequently used antibiotic as amoxicillin (including amoxicillin-clavulanate), a penicillin (Fig. 1A). There was specificity, or a differentiation, to antibiotic classes according to disease category. The top antibiotic class for respiratory tract infections (RTIs), urinary tract infections (UTIs), gastrointestinal tract infections (GIT), and dental cases, were, penicillins, sulfonamides, macrolides and sulfonamides, and macrolides, respectively (Fig. 1B).

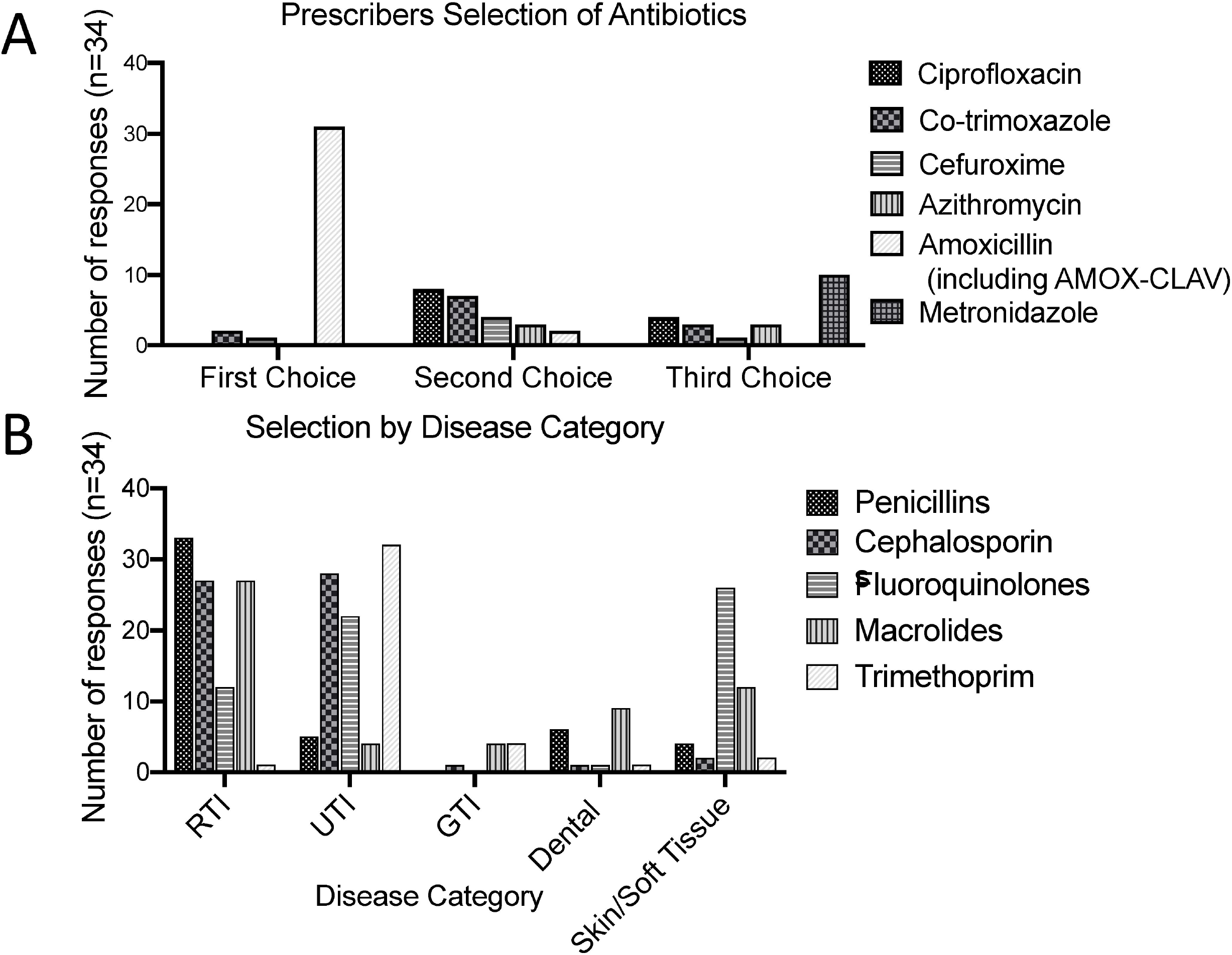

Physicians (65%, 22/34) report pressure to prescribe antibiotics. The reasons for this pressure, ranked from high to low, among participants who provided a reason, were the patients’ condition, 78% (14/18); perceived patient demand, 60% (12/20); fear of complications, 50% (8/16); and need to provide rapid relief, 47% (7/15).

The main considerations for prescribing antibiotics were the patients’ condition (94%, 32/34) and case clinical severity (62%, 21/34). The results from Antimicrobial Sensitivity Testing (AST) were a consideration only in about a third of respondents (29%, 10/34) – this being a Primary level facility without institutional AST.

Antibiotic choice was primarily determined by availability (87%, 26/30). Secondary considerations were inclusion in the Standard Treatment Guidelines (STG) or formulary (96%, 22/23); antibiotic spectrum (86%, 6/7); and patient symptoms (69%, 11/16). About one third (29%, 10/34) of respondents usually prescribe 2-3 antibiotics per antibiotic prescribing episode.

Generic prescribing, in line with existing protocols, was practiced by all prescribers (100%, 34/34). However, slightly over three quarters (76%, 26/34) also reported the practice of prescribing by brand names (likely outside the UNRWA system). The reasons for prescribing by brand was the perception that these are more efficacious (85%, 22/26) and/or of better quality (42%, 11/26) than generics.

The use of STG for prescribing was followed by 94% (32/34). There was limited AST capacity (Fig. 2). While about half (56%, n=34) responded that there was the capacity to perform AST, this was almost always (90%, n=34) off-site.

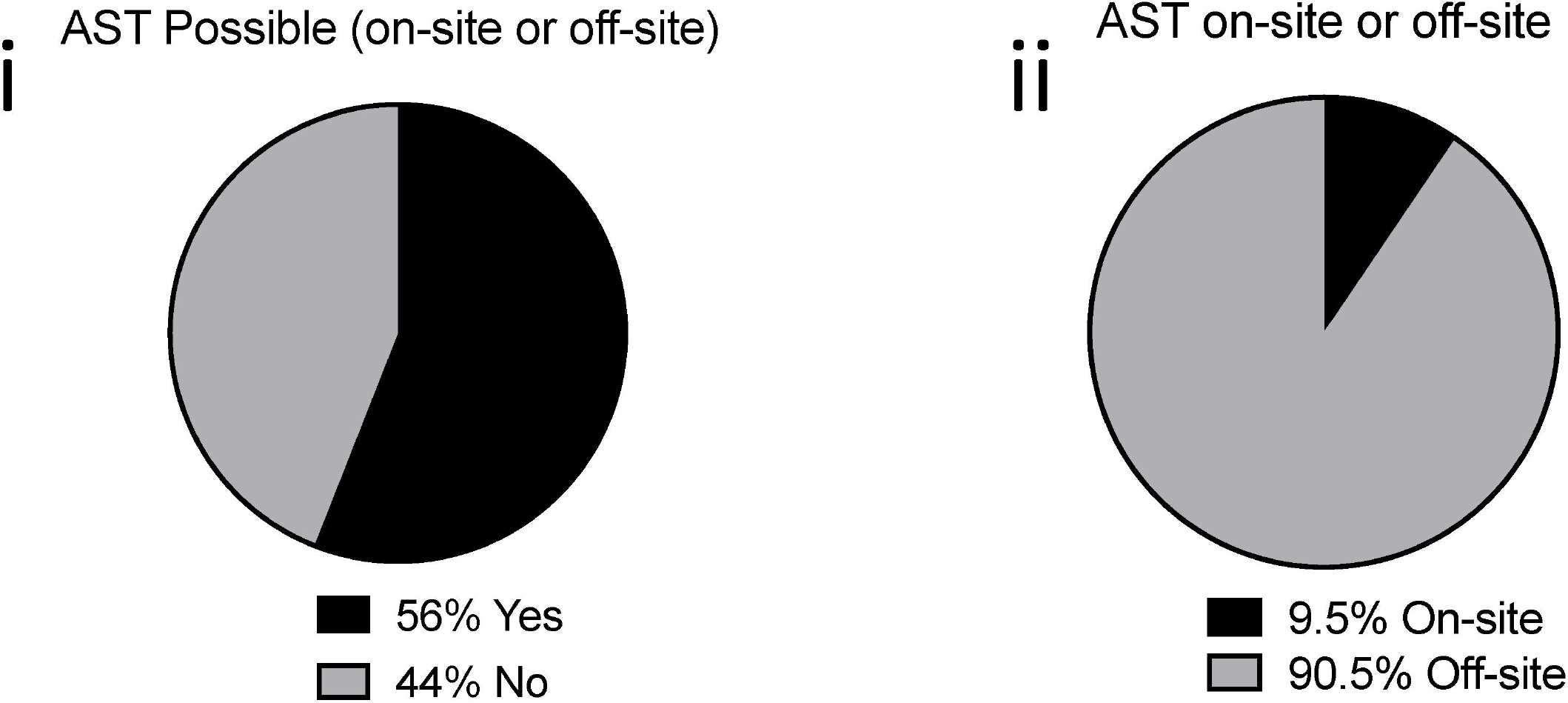

### Capability, opportunity and motivation for behavior change

#### Knowledge

There was a high real knowledge of AMR among participants. The great majority of prescribers correctly assessed the 7 knowledge statements (Table 2). All (100%, 34/34) knew that antibiotics were ineffective against viruses, including those causing the common cold and flu. Similarly, all (100, 34/34) had knowledge that every person treated with antibiotics is at an increased risk for developing an antibiotic resistant infection. The correct responses to the remaining 5 questions ranged from 82-97% (Table 2). The greatest relative knowledge gap was the fact that antibiotic resistance can spread from person to person (82% correct answer, n=34) ^17^.

**Table 2.** Real knowledge scores among prescribers at the UNRWA PHC, Jordan

|  |  | **TRUE,**<br>**n** | **FALSE,**<br>**n** | **Correct**<br>**answer** | **Correct**<br>**(%)** |
| --- | --- | --- | --- | --- | --- |
| **a** | Antibiotics are effective against viruses | 0 | 34 | FALSE | 100% |
| **b** | Antibiotics are effective against cold and flu | 0 | 34 | FALSE | 100% |
| **c** | Taking antibiotics has associated side effects such as diarrhea, colitis, or allergy | 34 | 0 | TRUE | 100% |
| **d** | Unnecessary use of antibiotics makes them become ineffective | 33 | 1 | TRUE | 97% |
| **e** | Antibiotic resistant bacteria can spread from person to person | 30 | 4 | TRUE | 88% |
| **f** | Every person treated with antibiotics is at an increased risk of antibiotic resistant infection | 34 | 0 | TRUE | 100% |
| **g** | Healthy people can carry antibiotic resistant bacteria | 32 | 2 | TRUE | 94% |
| Mean/aggregate real knowledge score |  |  |  |  | 97% |

#### Awareness

There was good awareness of the WHO 5 moments of hand-hygiene. All but one (97%, 33/34) could list the 5 moments. In contrast, there was poor awareness of the WHO AWaRe classification of antibiotics. Among participants, 41% (14/34) had heard of this classification.

#### Attitude

There was good attitude towards Infection Prevention and Control (hand hygiene) measures. All (100%, 34/34) affirmed the need to perform hand hygiene as often as recommended after contact with a patient or biological material.

#### Perceptions

Antimicrobial resistance was perceived to be a problem at both the PHC and country-level (Fig. 3). Four out of every five (79%, n=34) participants thought AMR was a problem at their PHC. Almost all 91% (31/34), thought AMR to be a problem in Jordan.

Antimicrobial resistance was perceived as a high or very high public health challenge in Jordan. Most (88%, 30/34) rated the AMR challenge as either very high or high. Refugees were perceived by 85% of respondents (n=34) to have a higher risk of AMR.

Increasing awareness among patients was perceived by almost all (94%, 32/34) prescribers as a solution to the problem of AMR in Jordan. Other top suggestions were enforcing prescription laws (85%, 29/34 of prescribers), increased AST (47%, 16/34); and training (38%, 13/34).

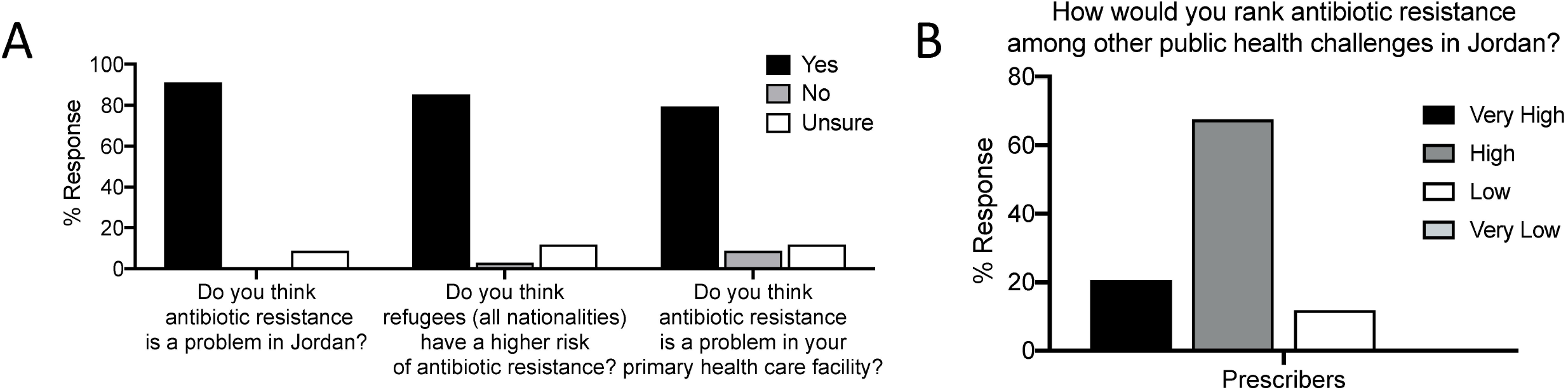

#### Opportunity

The aggregate opportunity score was 88%, indicating good opportunity for rational prescribing (Table 3).

**Table 3.** Opportunity to improve prescribing behavior

|  |  | Strongly<br>Disagree | Disagree | Neither<br>disagree<br>nor<br>agree | Agree | Strongly<br>agree | Total | Agree |
| --- | --- | --- | --- | --- | --- | --- | --- | --- |
| **a** | I have easy access to guidelines I need on managing infections | 2 |  |  | 21 | 11 | 34 | 94% |
| **b** | I am confident making antibiotic prescribing decisions |  | 2 | 1 | 21 | 10 | 34 | 91% |
| **c** | I have good opportunities to provide advice on prudent use to individuals (patients/public) |  | 2 | 2 | 19 | 11 | 34 | 88% |
| **d** | I have confidence in the antibiotic guidelines available to me |  | 3 | 1 | 20 | 10 | 34 | 88% |
| **e** | I consider antibiotic resistance when treating a patient | 2 |  | 3 | 17 | 12 | 34 | 85% |
| **f** | I have easy access to the materials I need to give advice on prudent antibiotic use and antibiotic resistance |  | 4 | 1 | 18 | 11 | 34 | 85% |
| **g** | I feel supported to not prescribe antibiotics when they are not necessary | 1 | 2 | 3 | 18 | 10 | 34 | 82% |
|  | Aggregate opportunity score |  |  |  |  |  |  | 88% |

Note: *Agree* is the combined proportion of “agree” and “strongly agree” responses expressed as percentage of all respondents (n=34).

#### Motivation

There was poor motivation among participants (Table 4). Less than 1 in 5 either believed that there was a correlation between their antibiotic prescribing and AMR (18%, 6/34), or that they have a key role in the control of AMR (15%, 5/34).

**Table 4.** Motivation for behavior change towards rational prescribing assessed by two statements and aggregate scores

|  |  | **Strongly**<br>**Disagree** | **Disagree** | **Neither**<br>**disagree**<br>**nor**<br>**agree** | **Agree** | **Strongly**<br>**agree** | **Agree** |
| --- | --- | --- | --- | --- | --- | --- | --- |
| **a** | There is a connection between my prescribing of antibiotics and emergence and spread of antibiotic resistant bacteria | 4 | 18 | 6 | 3 | 3 | 18% |
| **b** | I have a key role in helping control antibiotic resistance | 8 | 20 | 1 | 3 | 2 | 15% |
|  | Aggregate motivation |  |  |  |  |  | 16% |

Note: *Agree* is the combined proportion of “agree” and “strongly agree” responses expressed as percentage of all respondents (n=34).

#### Perceived solutions to improving prescribing

Among 14 options for improving prescribing, only 3 were “maybe acceptable” (Table 5). These were: availability of local guidelines (74%); availability of resistance data (74%); and educational sessions on prescribing (71%).

**Table 5.**
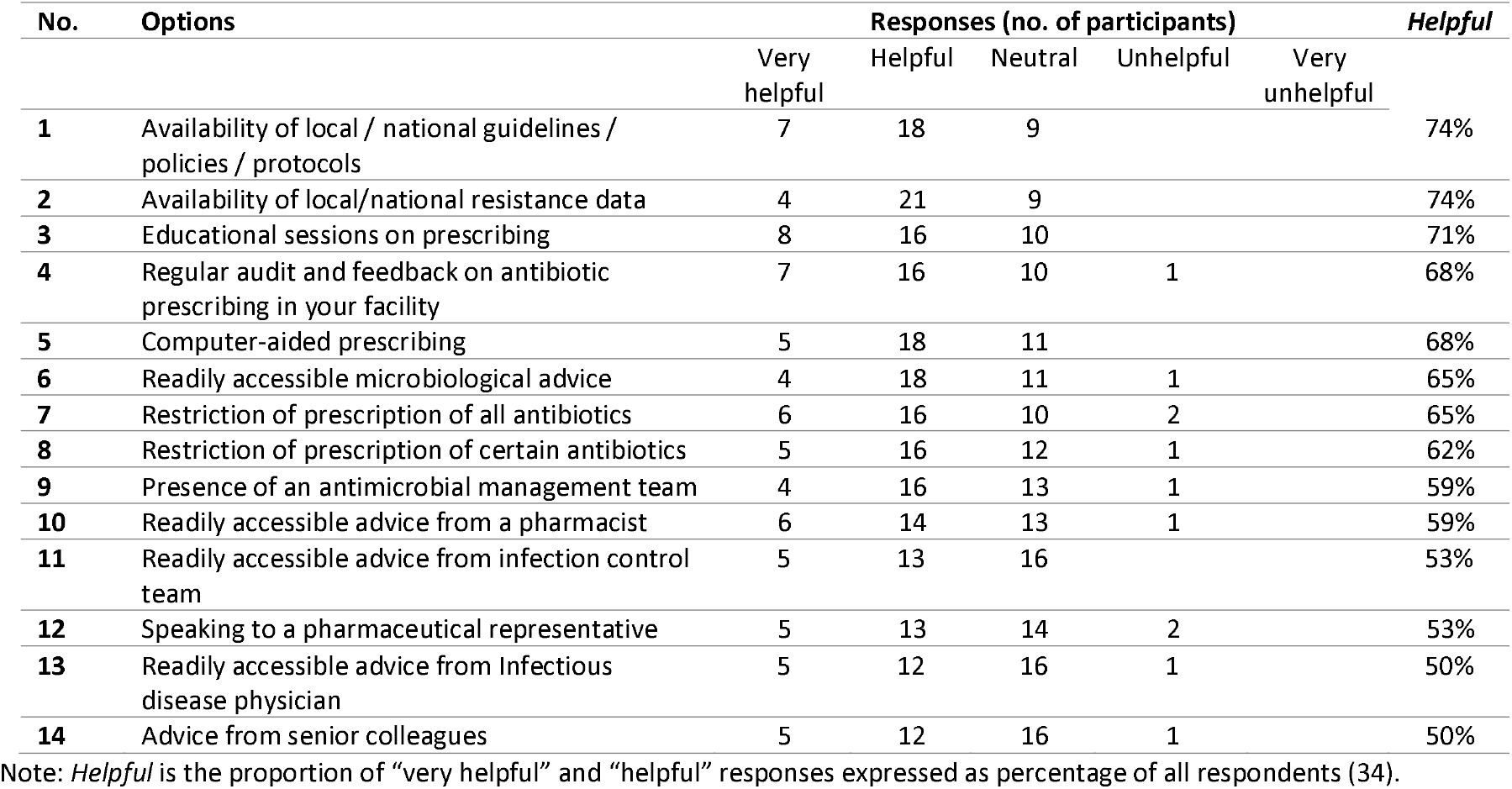
Options to improve antibiotic prescribing ranked by aggregate perceptions of “helpful” options among prescribers at UNRWA PHC, Jordan

## Discussion

Establishing needs through assessing behavioral, organizational and social barriers (including knowledge and awareness) and enablers provides evidence to design context-specific interventions aimed at curtailing the continuous spread of AMR. This study, conducted together with UNRWA, Jordan field of operation, provides a baseline evaluation to establish the need and parameters for intervention to designing, implementing and evaluation and monitoring of an effective AMS program at UNRWA.

In 2017, The Hashemite Republic of Jordan, in line with the global recommendation of the World Health Organization and the United Nations, formulated its National Action Plan to combat AMR for the period 2018-2022 ^18^. This plan, one of few that are fully costed, demonstrates the political commitment at a national level to addressing AMR in Jordan. The goals in this plan include a 30% reduction in antimicrobial use in animals, a 20% reduction in use in humans, and a 40% increase in public knowledge on AMR and awareness of appropriate use of antimicrobials. These goals are operationalized in two strategic objectives to improve knowledge on AMR and optimize use through AMS. These goals and objectives are shared by UNRWA.

Similarly, the UNRWA health department has a stated policy to maintain antibiotic prescription rates to less than 25%, reflecting earlier WHO recommendations on “optimum” antimicrobial medicines use/stewardship. In 2020, the Jordan field office reported a 16.6% antibiotic prescription rate, down from 21.3% in 2019, largely due to the impact of COVID-19 leading to a decrease in outpatient consultations and rational medicines prescription ^19,20^.

There was a high level of awareness of AMR among prescribers. By far a majority of participants rated AMR to be a challenge in Jordan as well as in their PHC. In addition, participants were of the opinion that refugees are at a higher risk of AMR than the general population. A few studies seem to suggest high carriage of AMR among the forcibly displaced probably as a result of certain socioeconomic factors ^21,22^

Overall, measures for antibiotic prescribing align with the WHO/INRUD recommendations. Most prescribers prescribe only 1 antibiotic per patient encounter or consultation; there is generic prescribing; and the formulary or STG guides antibiotic selection. The UNRWA field office has an electronic system that uses generic names and allows prescribing based only on the included generic medicines as derived or selected from the WHO Essential Medicines List.

The antibiotic prescribing pattern at UNRWA’s PHC Jordan field seem to mirror the prescribing pattern in the Middle East/North Africa (MENA) geographical region, with amoxicillin (including amoxicillin/clavulanate) being the most commonly prescribed antibiotic in Yemen, as well as in other states ^15^. In Jordan, this is an established trend. A 2007 study by Al-Niemat et al, found amoxicillin to be the most frequently prescribed ^23^. These results also agree broadly with a similar study conducted earlier in 2014 across five tertiary or referral outpatient public hospital pharmacies in Jordan which found amoxicillin to be the most prescribed antibiotic, followed by ciprofloxacin ^24^. Current national level antimicrobial consumption records also show amoxicillin and amoxicillin/clavulanate to be the most frequently consumed antibiotics in Jordan, accounting for 47% of total antibiotic consumption in Defined Daily Doses per 10, 000 population (DID) in 2019 and 32% in 2020 ^25^. The fall in 2020 was attributed to both COVID-19 and a simulated increase in other classes of antibiotics, compared to amoxicillin and the clavulanate.

The implications of this use pattern of amoxicillin on AMR, or clinical effectiveness, is not known. There is no information on the Global Antimicrobial Sensitivity Surveillance System (GLASS) for amoxicillin resistance. However, studies indicate very high levels of resistance (>80%) to this antibiotic in cases of Urinary Tract Infections, including those caused by Extended Spectrum Beta-Lactamase (ESBL) producing bacteria, in hospitalized children for the period 2012-2017 ^26^. A more recent, smaller, study found P aeruginosa to be 100% resistant to amoxicillin ^27^.

Antimicrobial prescribing, in practice, is a multi-layer exercise whereby the prescriber may have to balance certain considerations to resolve a case. These considerations are largely context specific. In this study, we found that physicians prioritized the condition of the patient in prescribing with the majority considering the condition of the patient in almost all cases, and or the clinical severity.

In line with common global practice, though, antibiotics were prescribed empirically, with limited considerations for AST. The apparent low prioritization of AST by prescribers is similar to another study in Yemen, though in this Jordan study, a higher proportion of prescribers rated as important the use of AST to guide rational prescribing than in Yemen, suggesting a higher level of awareness ^28^. The UNRWA PHC, it is to be noted, do not provide for AST services, being functionally out-patient level facilities. While there are no institutional AST services, some patients may go to private sector to do AST (as the Jordan field patients can access health services from both public and private sectors).

However, there may be a need to introduce AST or rapid diagnostic device services at the PHC to account for resistance rates. For example, results from GLASS, 2018, show that co-trimoxazole, the most commonly prescribed antibiotic for UTIs in this study would not be effective in 53% of cases. One study on antimicrobial use and resistance in a tertiary hospital in Jordan found an overall resistance prevalence of 26% across a panel of antibiotics including amoxicillin and the others commonly prescribed in this study in 33 patients receiving an antibiotic for a range of infections ^29^.

The use of a formulary, STG, or Essential Medicines List (EML) to guide antibiotic prescription is a AMS recommendation. This study suggests that prescribers follow this guideline in largely making decisions on choice of antibiotic to prescribe from the guidelines. Indeed, the UNRWA PHCs use an electronic prescribing and dispensing system that prevents prescribing outside this system. However, as the illustration with amoxicillin use and resistance shows, there is need for underlying dynamic resistance data to better guide antibiotic choice.

## Conclusion

This study assessed the knowledge, attitude and practices of prescribers at the UNRWA primary healthcare facilities in Jordan. There was a high knowledge of AMR and good opportunities, but poor motivation for rational prescribing or behavioral change. There is a clinical need for antimicrobial resistance data to promote rational antibiotic prescribing.

## Data Availability

All data produced in the present work are contained in the manuscript

## Acknowledgments

This work was made possible by support from the Boston University Social Innovation on Drug Resistance (SIDR) Postdoctoral Program to ESFO and the U.S. Pharmacopeial Convention (USP) Quality Institute to CC.

## Disclosures regarding real or perceived conflicts of interest

None to declare.

## Co-Author Contact Information

1. Albeik S; Department of health, headquarters, Amman, Jordan;
2. Ching C; Department of Biomedical Engineering, College of Engineering, Boston University, Boston, United States of America;
3. Hussein R; Department of Biomedical Engineering, College of Engineering, Boston University, Boston, United States of America;
4. Mousa A; Department of health, headquarters, Amman, Jordan;
5. Horino M; Department of health, headquarters, Amman, Jordan;
6. Naqa R; Department of health, Jordan Field office, Amman, Jordan;
7. Elayyan M; Department of health, Jordan Field office, Amman, Jordan;
8. Saadeh R; Department of health, headquarters, Amman, Jordan;
9. Zaman MH; Department of Biomedical Engineering, College of Engineering, Boston University, Boston, United States of America;

