## Supplement for "A survey assessing antimicrobial prescribing at UNRWA primary health care centers in Jordan"

Supplement 1. Survey_prescribers_UNRWA_Jordan_Jan24_2021

Background I: Antibiotic prescribing and patient characteristics

1) **Have you prescribed an antibiotic within the past 6 months?**

- Yes
- No

2) **How many patients do you see on a daily basis?**

- Less than 25
- Between 25-49
- Between 50-100
- Over 100

3) **Among the patients you see, approximately what percentage are children under 12 years old?**

- less than 10%
- between 10-24%
- between 25-50%
- over 50%

4) **Among the patients you see, approximately what percentage are Refugees (all nationalities)?**

- less than 25%
- between 25-49%
- between 50-75%
- Over 75%

5) **How many prescriptions for antibiotics have you written in the last 7 days?**

- Less than 50
- Between 50-99
- Between 100-149
- Over 150

Part One A: AMR Capacity

6) **To what extent do you agree or disagree with each of the following statements:**

|  | Strongly Disagree | | Disagree | Neither disagree nor agree | Agree | Strongly agree |
| --- | --- | --- | --- | --- | --- | --- |
| - I know what antibiotic resistance is | ☐_1_ | | ☐_2_ | ☐_3_ | ☐_4_ | ☐_5_ |
| - I know what information to give to individuals about prudent use of antibiotics and antibiotic resistance | ☐_1_ | | ☐_2_ | ☐_3_ | ☐_4_ | ☐_5_ |
| - I know sufficient knowledge about how to use antibiotics appropriately for my current practice | ☐_1_ | | ☐_2_ | ☐_3_ | ☐_4_ | ☐_5_ |

7) **Please answer whether you believe each of these statements to be True or False:**

|  | True | | | False |
| --- | --- | --- | --- | --- |
| - Antibiotics are effective against viruses | ☐ | | | ☐ |
| - Antibiotics are effective against cold and flu | ☐ | | | ☐ |
| - Unnecessary use of antibiotics makes them become ineffective | ☐ | | | ☐ |
| - Taking antibiotics has associated side effects such as diarrhea, colitis, or allergy | | ☐ | | ☐ |
| - Every person treated with antibiotics is at an increased risk of antibiotic-resistant infection | | ☐ | | ☐ |
| - Antibiotic resistant bacteria can spread from person to person | | ☐ | | ☐ |
| - Healthy people can carry antibiotic resistant bacteria | | ☐ | | ☐ |

Part One B: Perceptions of AMR

8) **Do you think antibiotic resistance is a problem in Jordan?**

- Yes
- No
- Unsure

9) **Do you think Refugees have a higher risk of antibiotic resistance than the general population?**

- Yes
- No
- Unsure

10) **Do you think antibiotic resistance is a problem in your primary health care facility?**

- Yes
- No
- Unsure

11) **How do you suggest the problem of antibiotic resistance can be tackled in Jordan? (Choose ALL that apply)**

- Increase testing for antibiotic susceptibility
- Increase awareness among patients
- Enforce prescription laws for the dispensing of antibiotics
- Improved training of doctors and pharmacists
- Other_______________________________________________________________________

Part three: Opportunity

12) **To what extent do you agree or disagree with each of the following statements:**

|  | Strongly Disagree | | | | Disagree | | Neither disagree nor agree | | Agree | | Strongly agree | |
| --- | --- | --- | --- | --- | --- | --- | --- | --- | --- | --- | --- | --- |
| - I have easy access to guidelines I need on managing infections | ☐_1_ | | | | ☐_2_ | | ☐_3_ | | ☐_4_ | | ☐_5_ | |
| - I have easy access to the materials I need to give advice on prudent antibiotic use and antibiotic resistance | ☐_1_ | | | | ☐_2_ | | ☐_3_ | | ☐_4_ | | ☐_5_ | |
| - I have good opportunities to provide advice on prudent use to individuals (patients/public) | ☐_1_ | | | | ☐_2_ | | ☐_3_ | | ☐_4_ | | ☐_5_ | |
| - I am confident making antibiotic prescribing decisions | | ☐_1_ | | | | ☐_2_ | | ☐_3_ | | ☐_4_ | | ☐_5_ |
| - I have confidence in the antibiotic guidelines available to me | | ☐_1_ | | | | ☐_2_ | | ☐_3_ | | ☐_4_ | | ☐_5_ |
| - I consider antibiotic resistance when treating a patient | | ☐_1_ | | | | ☐_2_ | | ☐_3_ | | ☐_4_ | | ☐_5_ |
| - I feel supported to not prescribe antibiotics when they are not necessary | | ☐_1_ | | | | ☐_2_ | | ☐_3_ | | ☐_4_ | | ☐_5_ |

Part four: Motivation

13) **To what extent do you agree or disagree with the following statements:**

|  | Strongly Disagree | | Disagree | Neither disagree nor agree | Agree | Strongly agree |
| --- | --- | --- | --- | --- | --- | --- |
| - There is a connection between my prescribing of antibiotics and emergence and spread of antibiotic resistant bacteria | ☐_1_ | | ☐_2_ | ☐_3_ | ☐_4_ | ☐_5_ |
| - I have a key role in helping control antibiotic resistance | ☐_1_ | | ☐_2_ | ☐_3_ | ☐_4_ | ☐_5_ |

Part two: One Health

14) **To what extent do you agree or disagree that the following environmental and animal health factors are important in contributing to antibiotic resistance in bacteria from humans?**

|  | Strongly Disagree | | Disagree | Neither disagree nor agree | Agree | Strongly agree |
| --- | --- | --- | --- | --- | --- | --- |
| - Environmental factors such as waste water in the environment | ☐_1_ | | ☐_2_ | ☐_3_ | ☐_4_ | ☐_5_ |
| - Excessive use of antibiotics in livestock and food production | ☐_1_ | | ☐_2_ | ☐_3_ | ☐_4_ | ☐_5_ |

Part five: Hand Hygiene

15) **Can you list the WHO’s five moments of hand hygiene?**

- Yes
- No
- Unsure

16) **Do you need to perform hand hygiene (as often as recommended) if you have used gloves in contact with patients or biological material?**

- Yes
- No
- Unsure

Part seven: AMS (Facilities, Services and Guidelines)

17) **In your facility, do you follow Standard Treatment Guidelines (or a guide specific to prescribing antibiotics)?**

- Yes
- No

18) **Does your facility have a Drug/Pharmacy and Therapeutic Committee (DTC/PTC)?**

- Yes
- No

19) **a. Is there a Healthcare Facility (or hospital) Formulary or an Essential Medicine List?**

- Yes
- No

**b. If yes, which (Please, check all that apply)** ?

- Hospital (UNRWA, or other standardized) Formulary
- Hospital or National Essential Medicine List/Rational Drug List

20) **a**. **Is it possible to perform laboratory antibiotic susceptibility testing on-site or off-site?**

- Yes
- No
- Not applicable

**b. If yes, is it on-site or off-site?**

- On-site
- Off-site

Part eight: Antibiotic prescribing; rationale and patterns

21) **How many patients, on average, do you see in a month?**

- 0 – 99
- 100 – 199
- 200 – 299
- 300 or more

-

22) **How often do you see treatment failure within a month (no response to therapy, switch to new antibiotic, or addition of new antibiotic)?**

- Less than 10%
- Between 11 – 25%
- Between 26 – 50%
- 50% or more

23) **a. Are** **antibiotic susceptibility tests performed before antibiotic selection?**

- Yes
- No

**b**. **If not, why? (Please, check all that apply)**

- Availability
- Time pressure
- Other. Please specify_____________________________________________

**c. If yes, how often?**

- Always
- Frequently
- Infrequently

**d. If yes, in what cases are antibiotic susceptibility tests usually used? (List up to three)**

______________________________________________________________________________

______________________________________________________________________________
______________________________________________________________________________

24) **Please select the top three antibiotics that you usually prescribe?**

|  | First Choice | Second Choice | Third Choice |
| --- | --- | --- | --- |
| - Amoxicillin (including amoxicillin-clavulanate) | ☐ | ☐ | ☐ |
| - Azithromycin | ☐ | ☐ | ☐ |
| - Cefuroxime | ☐ | ☐ | ☐ |
| - Ciprofloxacin | ☐ | ☐ | ☐ |
| - Co-trimoxazole | ☐ | ☐ | ☐ |
| - Metronidazole | ☐ | ☐ | ☐ |
| - Other: (Please list) _____________________ | ☐ | ☐ | ☐ |
| - Other: (Please list) _____________________ | ☐ | ☐ | ☐ |
| - Other: (Please list) _____________________ | ☐ | ☐ | ☐ |

25) **What is your main consideration for prescribing antibiotics?** **(Please, specify all that apply)**

- Antibiotic susceptibility testing information
- Symptoms of patient
- Clinical severity of case
- Other: Please, specify __________________________________________________________

26) **What are the top three things you considered for antibiotic CHOICE when you prescribe?**

|  | First | Second | Third |
| --- | --- | --- | --- |
| - Availability of antibiotic in the facility | ☐ | ☐ | ☐ |
| - Antibiotic susceptibility testing information | ☐ | ☐ | ☐ |
| - Broad-spectrum nature of antibiotic | ☐ | ☐ | ☐ |
| - Symptoms of patient | ☐ | ☐ | ☐ |
| - Other: (Please list) _____________________ | ☐ | ☐ | ☐ |
| - Other: (Please list) _____________________ | ☐ | ☐ | ☐ |
| - Other: (Please list) _____________________ | ☐ | ☐ | ☐ |

27) **When you prescribe antibiotics for a patient, do you usually prescribe? [Combination products count as 1]**

- 1
- 2-3
- >3

28) **Please indicate which GROUPs are MOST FREQUENTLY prescribed for the following infections?**

|  | RTI | | UTI | | GIT Diseases | | Dental Infections | | Skin & Soft Tissue infections | |
| --- | --- | --- | --- | --- | --- | --- | --- | --- | --- | --- |
| - Penicillins | ☐_1_ | | ☐_2_ | | ☐_3_ | | ☐_4_ | | ☐_5_ | |
| - Cephalosporins | ☐_1_ | | ☐_2_ | | ☐_3_ | | ☐_4_ | | ☐_5_ | |
| - Fluoroquinolones | ☐_1_ | | ☐_2_ | | ☐_3_ | | ☐_4_ | | ☐_5_ | |
| - Macrolides | | ☐_1_ | | ☐_2_ | | ☐_3_ | | ☐_4_ | | ☐_5_ |
| - Trimethoprim | | ☐_1_ | | ☐_2_ | | ☐_3_ | | ☐_4_ | | ☐_5_ |
| - Others (please specify):_________ | | ☐_1_ | | ☐_2_ | | ☐_3_ | | ☐_4_ | | ☐_5_ |

29) **Do you feel pressured to prescribe antibiotics?**

- Yes.
- No

30) **If yes, are any of these reasons that you feel pressured to prescribed antibiotics:**

|  | Strongly Disagree | | Disagree | | Neither disagree nor agree | | Agree | | Strongly agree | |
| --- | --- | --- | --- | --- | --- | --- | --- | --- | --- | --- |
| - Patient demand | ☐_1_ | | ☐_2_ | | ☐_3_ | | ☐_4_ | | ☐_5_ | |
| - Condition of patient | ☐_1_ | | ☐_2_ | | ☐_3_ | | ☐_4_ | | ☐_5_ | |
| - Fear of complications | ☐_1_ | | ☐_2_ | | ☐_3_ | | ☐_4_ | | ☐_5_ | |
| - Need to provide rapid relief | | ☐_1_ | | ☐_2_ | | ☐_3_ | | ☐_4_ | | ☐_5_ |
| - Other: (Please list) ________________ | | ☐_1_ | | ☐_2_ | | ☐_3_ | | ☐_4_ | | ☐_5_ |

31) **Do you usually prescribe antibiotics by generic or brand name?**

- Generic
- Brand

32) **If you prescribe by brand, what are your reasons for doing so? (Please, specify all that apply)**

- Clinical efficacy (high bacterial resistance to generics)
- Better quality
- Other (please specify) ____________________________________________________________

33) **What measures do you think would be the most helpful in improving antibiotic prescribing?**

|  | | Very helpful | | Helpful | Neutral | Unhelpful | Very  unhelpful |
| --- | --- | --- | --- | --- | --- | --- | --- |
| - Educational sessions on prescribing | | ☐_1_ | | ☐_2_ | ☐_3_ | ☐_4_ | ☐_5_ |
| - Availability of local / national   guidelines / policies / protocols | | ☐_1_ | | ☐_2_ | ☐_3_ | ☐_4_ | ☐_5_ |
| - Availability of local/national resistance data | | ☐_1_ | | ☐_2_ | ☐_3_ | ☐_4_ | ☐_5_ |
| - Computer-aided prescribing | | ☐_1_ | | ☐_2_ | ☐_3_ | ☐_4_ | ☐_5_ |
| - Presence of an antimicrobial management team | | ☐_1_ | | ☐_2_ | ☐_3_ | ☐_4_ | ☐_5_ |
| - Readily accessible microbiological advice | | ☐_1_ | | ☐_2_ | ☐_3_ | ☐_4_ | ☐_5_ |
| - Readily accessible advice from Infectious Disease physician | | ☐_1_ | | ☐_2_ | ☐_3_ | ☐_4_ | ☐_5_ |
| - Readily accessible advice from a pharmacist | ☐_1_ | | | ☐_2_ | ☐_3_ | ☐_4_ | ☐_5_ |
| - Readily accessible advice from infection control team | ☐_1_ | | | ☐_2_ | ☐_3_ | ☐_4_ | ☐_5_ |
| - Advice from senior colleagues | ☐_1_ | | | ☐_2_ | ☐_3_ | ☐_4_ | ☐_5_ |
| - Speaking to a pharmaceutical representative | ☐_1_ | | | ☐_2_ | ☐_3_ | ☐_4_ | ☐_5_ |
| - Restriction of prescription of certain antibiotics | ☐_1_ | | | ☐_2_ | ☐_3_ | ☐_4_ | ☐_5_ |
| - Restriction of prescription of **all** antibiotics | ☐_1_ | | | ☐_2_ | ☐_3_ | ☐_4_ | ☐_5_ |
| - Regular audit and feedback on antibiotic prescribing in your PHC | ☐_1_ | | | ☐_2_ | ☐_3_ | ☐_4_ | ☐_5_ |

34) **To optimize the use of antibiotics, the WHO has classified antibiotics as Access, Watch, & Reserve.**
a. **Have you heard about these categories?**

- Yes
- No

b. **If yes, do they affect your prescribing?**

- Yes
- No

35) **How many years of practice experience do you have?**

- Less than 1 year
- 1 – 5 years
- 6 – 10 years
- 11 – 15 years
- 15 years or more

36) **What is your specialty?**

- Medicine
- Surgery
- Pediatrics
- Anesthetics
- Obstetrics / Gynecology
- Psychiatry
- Other

Part nine: Additional (Optional)

37) **Do you have additional comments you find relevant?**

    ______________________________________________________________________________
    ______________________________________________________________________________

______________________________________________________________________________

______________________________________________________________________________
______________________________________________________________________________

**Thank you very much for taking part in this survey!**

Supplement 2 Survey results.

|  |  | n | % |
| --- | --- | --- | --- |
| 1 | Consent |  |  |
|  | Yes | 34 | 92% |
|  | No | 3 | 8% |
| 2 | Prescribed antibiotic past 6 months |  |  |
|  | Yes | 34 | 100% |
|  | No | 0 |  |
| 3 | Patients, daily load |  |  |
|  | <25 | 1 | 3% |
|  | 25-49 | 1 | 3% |
|  | 50-100 | 28 | 82% |
|  | >100 | 4 | 12% |
| 4 | Cliente under 12 |  |  |
|  | <10% | 3 | 9% |
|  | 10-<25% | 26 | 76% |
|  | 25-50% | 5 | 15% |
|  | >50% |  |  |
| 5 | a. Palestinian refugees as proportion of cliente |  |  |
|  | <25% | 1 | 3% |
|  | 50-75% | 1 | 3% |
|  | >75% | 32 | 94% |
|  | b. Palestine refugees from Jordan |  |  |
|  | <25%  25-49% |  |  |
|  | 50-75% |  |  |
|  | >75% |  |  |
|  | c. Palestine refugees from Syria |  |  |
|  | <25% |  |  |
|  | 50-75% |  |  |
|  | >75% |  |  |
|  | d. Ex-Gazan refugees |  |  |
|  | <25% |  |  |
|  | 50-75% |  |  |
|  | >75% |  |  |
|  | e. Other refugees |  |  |
|  | <25% |  |  |
|  | 50-75% |  |  |
|  | >75% |  |  |
| 6 | Antibiotic prescriptions last 7 days |  |  |
|  | <50 | 7 | 21% |
|  | 50-99 | 12 | 35% |
|  | 100-149 | 10 | 29% |
|  | >150 | 5 | 15% |
| 7 | AMR a problem in Jordan |  |  |
|  | Yes | 31 | 91% |
|  | No | 3 | 9% |
| 8 | Refugees have a higher risk of AMR |  |  |
|  | Yes | 29 | 85% |
|  | No | 1 | 3% |
|  | Unsure | 4 | 12% |
| 9 | AMR a problem at your PHC |  |  |
|  | Yes | 27 | 79% |
|  | No | 3 | 9% |
|  | Unsure | 4 | 12% |
| 10 | Tackling AMR |  |  |
|  | Increased testing | 16 | 47% |
|  | Increase patient awareness | 32 | 94% |
|  | Enforce prescription laws | 29 | 85% |
|  | Improved training for doctors and pharmacists | 13 | 38% |
| 11 | WHO's five moments hand hygeine |  |  |
|  | Yes | 33 | 97% |
|  | No |  |  |
|  | Unsure | 1 | 3% |
| 12 | Hand hygeine after using gloves |  |  |
|  | Yes | 33 | 97% |
|  | No |  |  |
|  | Unsure | 1 | 3% |
| 13 | Use of STG at facility |  |  |
|  | Yes | 33 | 97% |
|  | No |  |  |
|  | Unsure | 1 | 3% |
| 14 | DTC at facility |  |  |
|  | Yes | 20 | 74% |
|  | No | 6 | 22% |
| 15 | a. Hospital Formulary or EML |  |  |
|  | Yes | 24 | 89% |
|  | No | 3 | 11% |
|  | b. If Yes, which |  |  |
|  | Formulary | 9 | 33% |
|  | EML | 8 | 30% |
|  | Both | 5 | 19% |
| 16 | a. Possibility to perform AST onsite or offsite |  |  |
|  | Yes | 19 | 56% |
|  | No | 4 | 12% |
|  | Not applicable | 11 | 32% |
|  | b. If Yes, is it on-site or off-site |  |  |
|  | On-site | 2 | 11% |
|  | Off-site | 17 | 89% |
| 17 | Patients, montly load |  |  |
|  | 0-99 | 3 | 9% |
|  | 100-199 | 3 | 9% |
|  | 200-299 | 11 | 32% |
|  | >300 | 17 | 50% |
| 18 | Treatment failure |  |  |
|  | <10% | 7 | 21% |
|  | 11-25% | 22 | 65% |
|  | 26-50% | 5 | 15% |
|  | >50% |  |  |
| 19 | a. AST before antibiotic selection |  |  |
|  | Yes | 7 | 21% |
|  | No | 27 | 79% |
|  | b. If not, why? |  |  |
|  | Availability | 12 | 44% |
|  | Time pressure | 19 | 70% |
|  | Other |  |  |
|  | Finance | 2 | 7% |
|  | Very ill patient | 4 | 15% |
|  | c. If yes, how often are AST performed |  |  |
|  | Always |  |  |
|  | Frequently | 2 | 7% |
|  | Infrequently | 6 | 22% |
|  | d. If yes, cases when AST are used |  |  |
|  | UTI in pregnancy, wound sepsis, puerperal sepsis, post-abortion sepsis, persistent fever unresponsive to treatment, bronchopneumonia, meningitis, otitis media |  |  |
| 20 | No of antibiotics usually prescribed |  |  |
|  | One |  |  |
|  | Two-three | 10 | 37% |
|  | More than 3 |  |  |
| 21 | Pressure to prescribe antibiotics |  |  |
|  | Yes | 15 | 56% |
|  | No | 12 | 44% |
| 22 | a. Prescribing |  |  |
|  | Brand | 8 | 30% |
|  | Generic | 19 | 70% |
|  | b. Reasons for brand prescribing (select all that apply) |  |  |
|  | Clinical efficacy (high bacterial resistance to generics) | 5 | 19% |
|  | Better quality | 9 | 33% |
|  | Others [severity of the symptoms] |  |  |
| 23 | a. Heard about WHO AWARE categories |  |  |
|  | Yes | 4 | 15% |
|  | No | 23 | 85% |
|  | b. If yes, do they affect your prescribing behaviour |  |  |
|  | Yes | 2 | 7% |
|  | No | 25 | 93% |
| 24 | Years of practice experience |  |  |
|  | 1-5 years | 4 | 15% |
|  | 6-10 years | 10 | 37% |
|  | 11-15 years | 7 | 26% |
|  | >15 years | 6 | 22% |

Notes: The table excludes results presented within the main manuscript.
